# The Human Disharmony Loop: Demystifying Thoracic Outlet Syndrome

**DOI:** 10.64898/2025.12.03.25341566

**Authors:** Ketan Sharma, Jaicharan J. Iyengar, James Friedman

**Affiliations:** St. Luke’s Plastic and Reconstructive Surgery, Meridian, ID, 83642; Sutter Alpine Care Clinic, Stockton, CA, 95204

## Abstract

**Background:** Thoracic outlet syndrome (TOS) remains controversial, with contentious diagnostic criteria and morbid surgeries of questionable efficacy. Although considered neurovascular compression, many mysteries abound, including presence of neck and upper back pain, occipital headaches, scapular protraction, and higher prevalence in women. Previously, we described the Human Disharmony Loop (HDL), where the asymmetric lower trunk innervation to the pectoralis minor (PM) protracts the scapula and deforms its connections. The pathoanatomy includes narrowing the thoracic outlet and subacromial spaces, stretching the upper trapezius and rhomboids, and irritating the occipital nerves, generating occipital headaches, upper back and neck pain, shoulder weakness, and radiating neuropathy. We hypothesize that TOS is a scapular phenomenon produced after PM tightness, and hence these patients will benefit from PM tenotomy with infraclavicular plexus neurolysis (PM+ICN) alone.

**Methods:** TOS patients with 6-month minimum follow-up who met HDL diagnostic criteria were treated with PM+ICN. Outcomes included pain, shoulder abduction range of motion, headaches, scapular dyskinesis, secondary neurolysis.

**Results:** *N* = 144 patients were included. Median age was 50; 76% were female. Following PM+ICN, median pain decreased from 8.0/10 to 2.0/10, median shoulder abduction increased from 90 to 180 degrees, occipital headaches decreased from 83% to 1%. Scapular dyskinesis normalized from 99% protraction at rest to 94% no protraction (all *p*<0.05). 16% required secondary neurolysis with only 1% at the thoracic outlet. Median follow-up was 15 months.

**Conclusions:** In a large series, isolated PM+ICN normalized scapular mechanics and substantially reduced pain and improved function. These findings suggest that TOS may be an anatomic subset of the HDL: the PM is the true anatomic culprit. The HDL anatomically answers the mysteries of TOS including headaches, neck pain, scapular protraction, and higher prevalence in women. These historically challenging patients may benefit substantially from an effective and low morbid alternative, PM+ICN.

**Level of Evidence:** IV, therapeutic.

## Introduction

Thoracic outlet syndrome (TOS) remains a notoriously-challenging chronic pain syndrome of the upper limb.[1] First described in 1956[2], patients present with pain, numbness/tingling, weakness, and temperature abnormalities.[3, 4] Diagnosis relies on patient presentation, physical exam maneuvers, and ruling out competing pathologies.[5, 6] Fundamentally, the etiology is attributed to compression of the brachial neurovascular bundle in the interscalene, costoclavicular, and/or retrocoracoid regions.[7] TOS is categorized into venous (vTOS), arterial (aTOS), and neurogenic (nTOS) subtypes. Of these, nTOS constitutes the overwhelming majority but also carries the worst prognosis.[7–9] Surgery is typically advised after failure to improve despite 6 months of conservative therapy.[7] Surgical treatments include scalenectomy and neurolysis with or without first rib resection (SCN±FRR), via transaxillary, supraclavicular, or infraclavicular approaches, with some adding pectoralis minor (PM) tenotomy to address retrocoracoid compression.[10, 11] Recent advances include less invasive robotic and thoracoscopic techniques[10], with ongoing debate regarding the risk versus benefit of FRR.[12]

However, there are “few topics more controversial”[4], with many disputing TOS’s existence[3], for three principal reasons.[13] First, the diffuse and vague symptoms overlap with other syndromes, including cervical radiculopathy, peripheral neuropathy, and complex regional pain syndrome.[14, 15] Second, the diagnosis is not reproducible[5, 16–18]: physical exam tests suffer from unreliable specificity and sensitivity[19], while EMGs and imaging are mostly negative.[7, 20, 21] Third, current surgeries do not convincingly demonstrate efficacy[13], and even botulinum toxin injections into the scalene muscles, the alleged culprit, confers no benefit over placebo.[22] Surgeries can be morbid with complication rates nearing 20%[23], and exhibit lofty recurrence rates, as the initial symptom resolution gradually deteriorates.[12, 24]

Additional mysteries abound. Patients also exhibit symptoms that cannot be explained by brachial neurovascular compression, including neck, upper back, peri-trapezial, and peri-clavicular pain[12, 14, 25, 26], occipital headaches[27, 28], and chronically “slumped” postures with hunched shoulders.[4] The ipsilateral scapula is well-known to protract while reaching overhead.[29, 30] Explanations for this phenomenon tend to be convoluted or non-anatomic, such as “muscle imbalance”[4], over-compensation by the upper trapezius[29], and an isolated “Sunderland zero” lesion to the long thoracic nerve.[31] The latter is incongruent with the fact that the entire plexus is subject to compression. Furthermore, TOS occurs far more commonly in women[7], but the proposed rationales of hormones and differences in pain perception[32] remain unconvincing, if not condescending and patriarchal. While the supposed proximate cause is compression, a myriad of ultimate sources underlying the compression tend to be invoked, including variations in cervical rib and scalene muscle anatomy, anomalous fibro-fascial bands, repetitive trauma and strain injury, excessive overhead activity, postural changes, altered shoulder girdle mechanics, and scalene and PM spasm.[7, 8, 19, 26] However, these remain incredibly vague, and how they arise to begin with is left unexplained.

Thus, numerous considerable lines of evidence converge on the conclusion that the current anatomic understanding of TOS is incomplete at best.

Previously, we described the Human Disharmony Loop (HDL), a unifying clinical model of upper limb chronic pain.[33, 34] The scapula coordinates all function between the body (thorax) and arm (humerus).[35] The peri-scapular stabilizers control this motion, of which, the ventral PM uniquely carries lower trunk C8-T1 innervation. This neurologic asymmetry, likely an evolutionary sequelae of the transition from quadrupeds to bipeds[33], renders the human scapula dyskinetic in a self-reinforcing cycle. (Figure 1) Protraction then deforms all the scapula’s connections. One pathoanatomic consequence is to lower the acromion and clavicle and therefore narrow the thoracic outlet itself. Others include impinging the subacromial space, stretching the dorsal upper trapezius and rhomboids, tensioning the brachial plexus mainly the upper trunk, and irritating the occipital nerves to the posterior scalp. Hence, a single source – PM tightness – generates four clusters of symptoms: occipital headaches and neck stiffness, upper back tightness, shoulder weakness, and radiating numbness/tingling to the hand worse with overhead reach. Crucially, the HDL is diagnosed purely on history and physical exam. (Figure 2)

**Figure 1.**
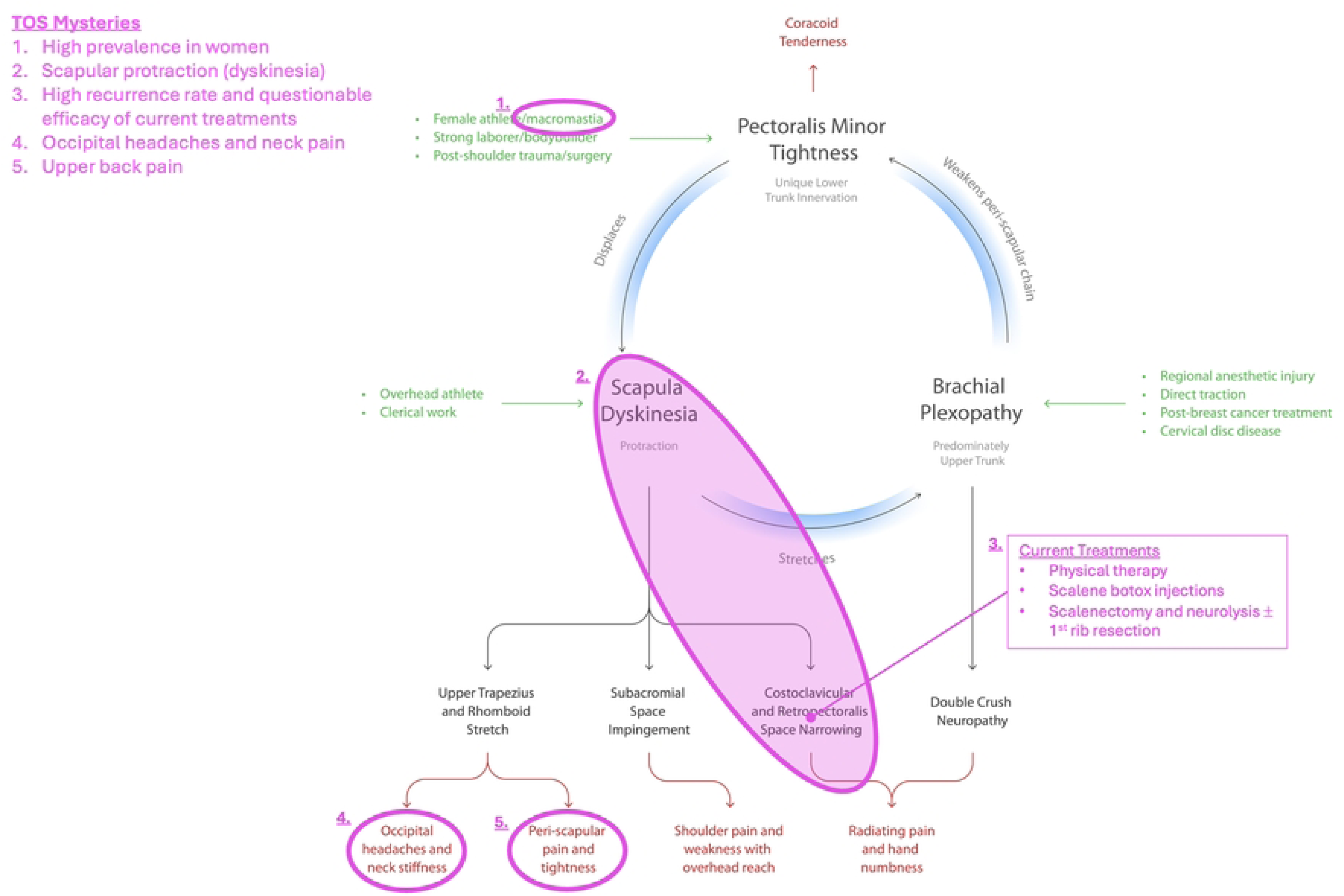
Thoracic Outlet Syndrome (TOS) as a Subset of the Human Disharmony Loop (HDL) TOS is the subset of the HDL where scapular protraction tugs the clavicle down via the AC joint, thereby narrowing the thoracic outlet and compressing the neurovascular bundle. The many mysteries of conventional TOS which cannot be explained by neurovascular compression are anatomically incorporated into the HDL, including (1) higher prevalence in women, (2) scapular protraction on exam, (3) inefficacy of current surgeries, (4) occipital headaches, and (5) upper back pain.

**Figure 2.**
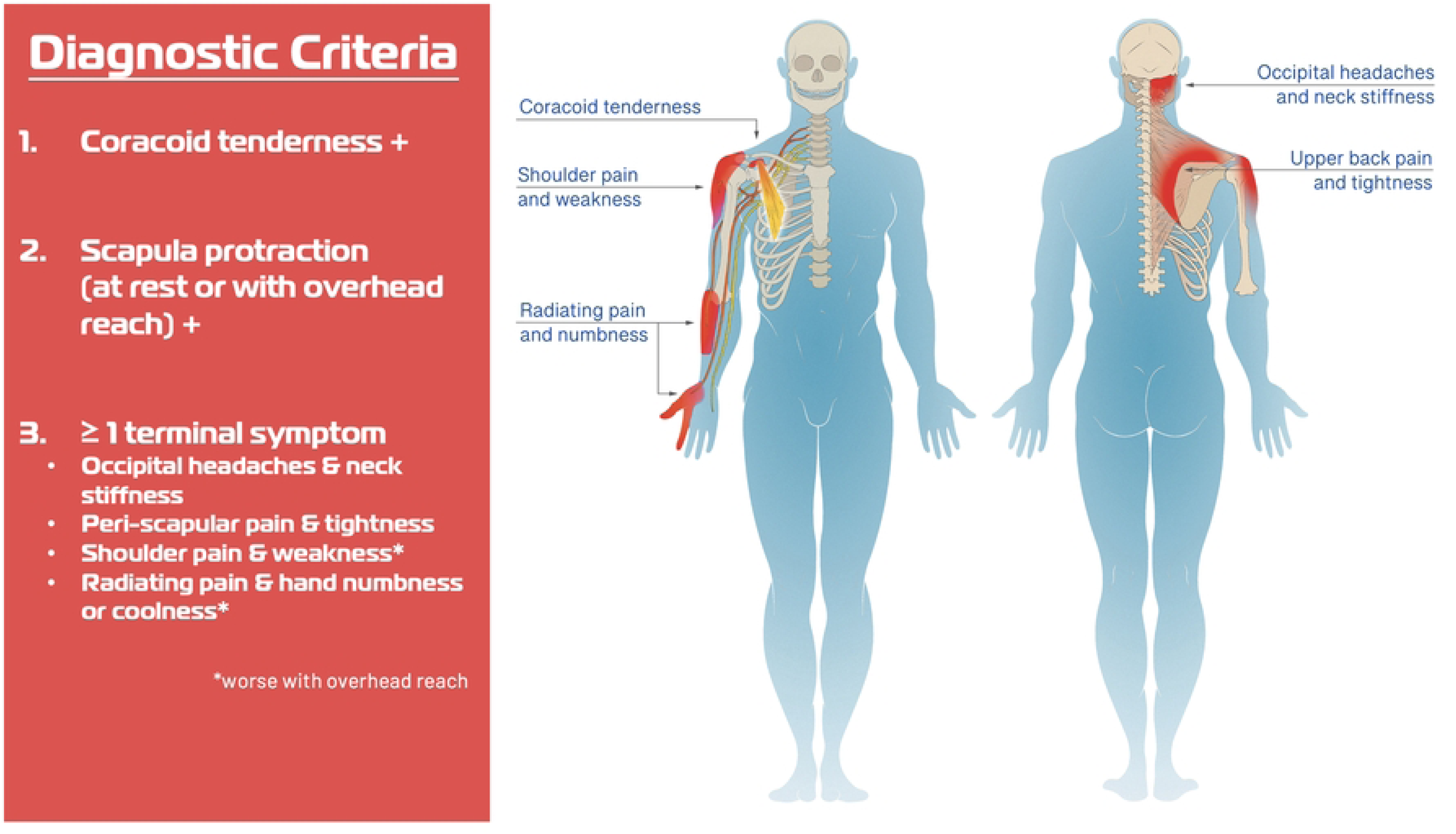
Human Disharmony Loop Diagnostic Criteria Diagnosis of the HDL is based on two anatomic and one symptomatic criterion and derives entirely from history and physical exam

In our first three reports[33, 34, 36], we noted that HDL patients with a history of TOS displayed significant improvements in pain, headaches, and motion following PM tenotomy with infraclavicular neurolysis (PM+ICN), including some who had failed prior TOS surgery. Furthermore, the HDL can anatomically explain the mysteries of TOS: scapular protraction, occipital headaches, neck and upper back pain, shoulder weakness, increased incidence in women, and inefficacy of current surgeries. These considerations suggest that TOS may be primarily a scapular phenomenon – a subset of the HDL. The neurovascular compression within the thoracic outlet may be secondary to the deforming pull of the PM, and therefore relieved by solely targeting the PM with proper rehabilitation to restore scapular retraction. Therefore, we hypothesize that PM+ICN alone will benefit TOS patients who meet HDL criteria, avoiding need for far more morbid SCN±FRR.

## Materials and Methods

This is a combined retrospective and prospective case series of consecutive patients treated at two sites. All were evaluated by a fellowship-trained board-certified hand, sports, or shoulder surgeon. Inclusion criteria included: age > 18 years, a diagnosis of TOS, and meeting HDL diagnostic criteria. Exclusion criteria included: follow-up < 6 months. Patients were diagnosed with HDL based on history and physical (Figure 2) and with TOS based on Society of Vascular Surgeons criteria.[6] All patients trialed at least 3 months of physical therapy (PT) before being offered surgery. At each visit, patients completed a self-reported pain questionnaire. Shoulder abduction range of motion (ROM) values were measured. Scapula dyskinesis was classified via physical exam into four stages: none (no protraction), dynamic (protraction with overhead reach only), static reversible (protraction at rest but manually reversible via the examiner), static irreversible (protraction at rest not manually reversible via the examiner). Neuropathy was diagnosed on exam via the scratch collapse test (SCT).[37] Patients were offered a preoperative medial coracoid injection for symptom relief, but a negative response did not rule out the HDL.[34, 36] Patients underwent open PM+ICN, followed by PT emphasizing upper trapezius and rhomboid strengthening to restore scapular retraction.[33] At 3 months, patients with persistent pain and/or weakness were offered secondary neurolysis for lingering neuropathic lesions on exam. Outcomes included pain, headaches, neuropathic lesions, and shoulder abduction ROM. Wilcoxon rank-sum and Chi-squared analysis compared continuous and categorical variables of interest, respectively. All analysis was performed using STATA v14.0. Institutional Review Board (IRB) approval was obtained. Informed consent was waived by the ethics committee as all data was anonymous and the study posed minimal risk to patients. The retrospective arm occurred from 1/13/23-10/10/24, and the prospective arm was from 10/11/24-2/2/25. The authors did not have access to information that could identify individual participants after data collection. This study conforms to the STROBE guidelines.

## Results

*N* = 144 patients were included. Subtypes were 98% neurogenic, 2% venous. Median age was 50. Sex was 24% male and 76% female. Prior history included 26% fibromyalgia, 40% cervical stenosis, 9% scapular dyskinesis, and 85% subacromial pain. Surgical history included 22% subacromial decompression with adjunct procedures, 7% total shoulder arthroplasty, 2% SCN+FRR, 19% cervical fusion, 31% carpal and/or cubital tunnel release. (Table 1) Median pre-operative pain was 8.0/10. Scapular dyskinesis was 1% dynamic, 41% static reversible, 58% static irreversible. Neuropathy at presentation was: 100% thoracic outlet, 72% suprascapular, 92% axillary, 68% radial, 25% ulnar, and 49% median. Median shoulder abduction was 90 degrees, and 83% endorsed occipital headaches. 105 patients accepted a medial coracoid injection, of which 88% endorsed symptom relief. Postoperatively, median pain decreased to 2.0/10. Scapular dyskinesis redistributed to 94% none, 5% dynamic, and 1% static reversible. Neuropathy decreased to 3% thoracic outlet, 1% suprascapular, 14% axillary, 16% radial, 20% cubital, and 16% carpal. Median shoulder abduction increased to 180 degrees, and headaches decreased to 1%. All changes were statistically significant (*p*<0.05). Median follow-up was 15 months. 16% required subsequent neurolysis at the following: 1% thoracic outlet, 5% quadrilateral space, 6% radial tunnel, 8% cubital tunnel, and 3% carpal tunnel. (Table 2)

**Table 1.** Patient Variables.

| Variable | *N* = 144 |
| --- | --- |
| Age<sup>1</sup> | 50 [39 – 65] |
| Sex | Male 34 (24%)<br>Female 110 (76%) |
| BMI<sup>1</sup> | 30 [26 – 34] |
| Concomitant Diagnosis |  |
| Fibromyalgia | 38 (2%) |
| Cervical disc disease | 58 (40%) |
| Scapular dyskinesis | 13 (9%) |
| Subacromial impingement | 123 (85%) |
| Surgical History |  |
| Subacromial decompression + adjunct procedures<sup>2</sup> | 32 (22%) |
| Reverse total shoulder arthroplasty | 10 (7%) |
| 1<sup>st</sup> rib resection + scalenectomy | 3 (2%) |
| Cervical spine fusion | 27 (19%) |
| Distal neurolysis (carpal, cubital) | 44 (31%) |
| Hand Dominance | Right 124 (86%)<br>Left 20 (14%) |
| Laterality | Right 93 (65%)<br>Left 51 (35%) |
| TOS Subtype | Neurogenic 141 (98%)<br>Venous 3 (2%) |
| Medial Coracoid Injection | Positive Response 105/119 (88%) |
| Follow-up (months)<sup>1</sup> | 15 [12-28] |

1. Median [inter-quartile range] 2. Adjunct procedures include rotator cuff repair, distal clavicle excision, biceps tenodesis, bursectomy and/or labral repair

**Table 2.**
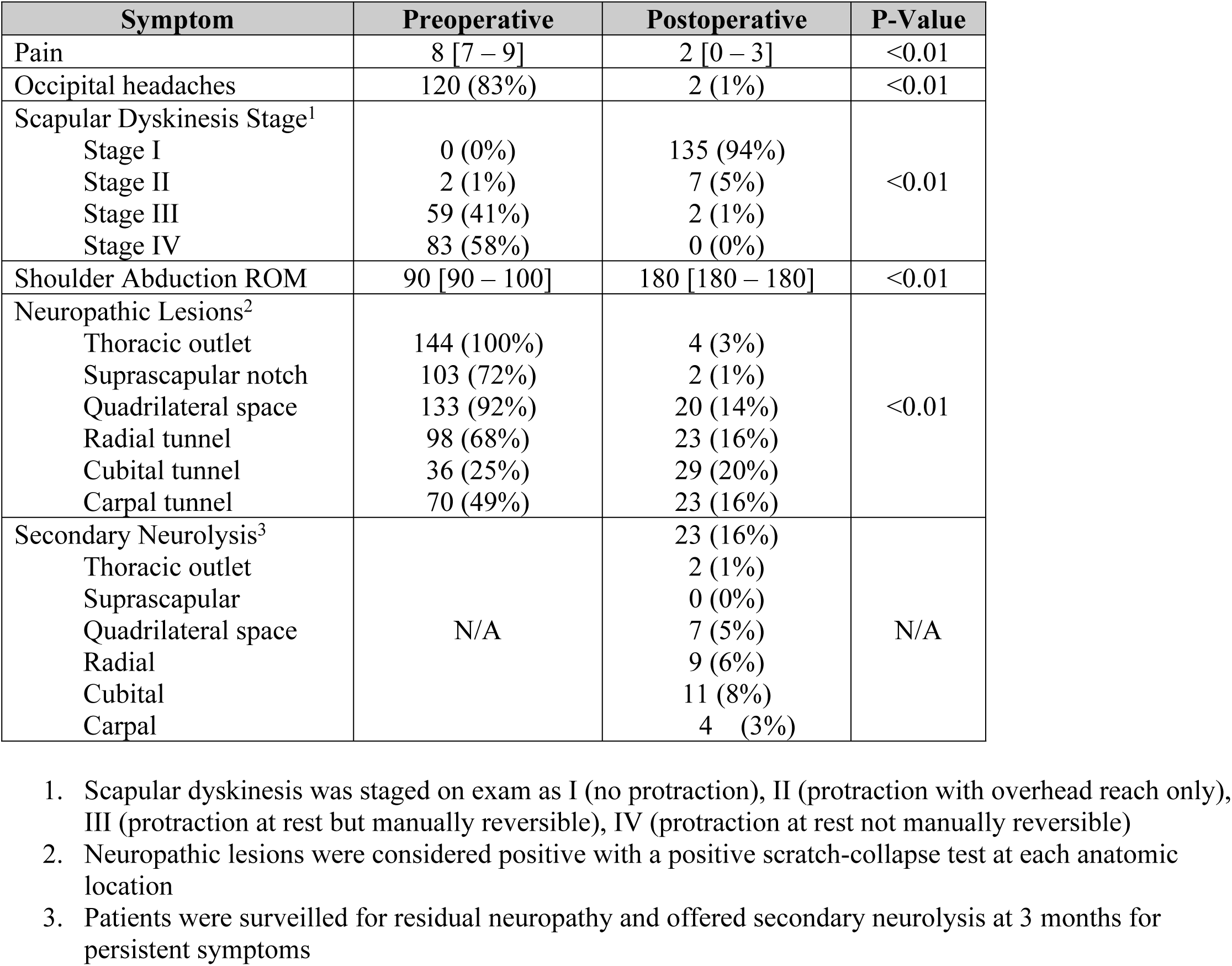
Clinical Outcomes.

## Discussion

In this study, in a large series of predominantly nTOS patients, PM+ICN normalized scapular mechanics and substantially reduced pain, neuropathy, and headaches, while restoring shoulder ROM. (Figure 3) A minority required subsequent neurolysis for full relief, but only 1% at the thoracic outlet itself, where current and often morbid surgeries target. Hence, we assert that TOS may be fundamentally a scapular phenomenon, a subset of the HDL. PM tightness protracts the scapula and deforms its connections[36], including pulling the clavicle down via the AC joint and narrowing the costoclavicular space, generating TOS. (Figure 4) Current surgeries particularly SCN±FRR target solely the focal brachial neurovascular compression but ultimately miss the root anatomic source, explaining their inefficacy.

**Figure 3.**
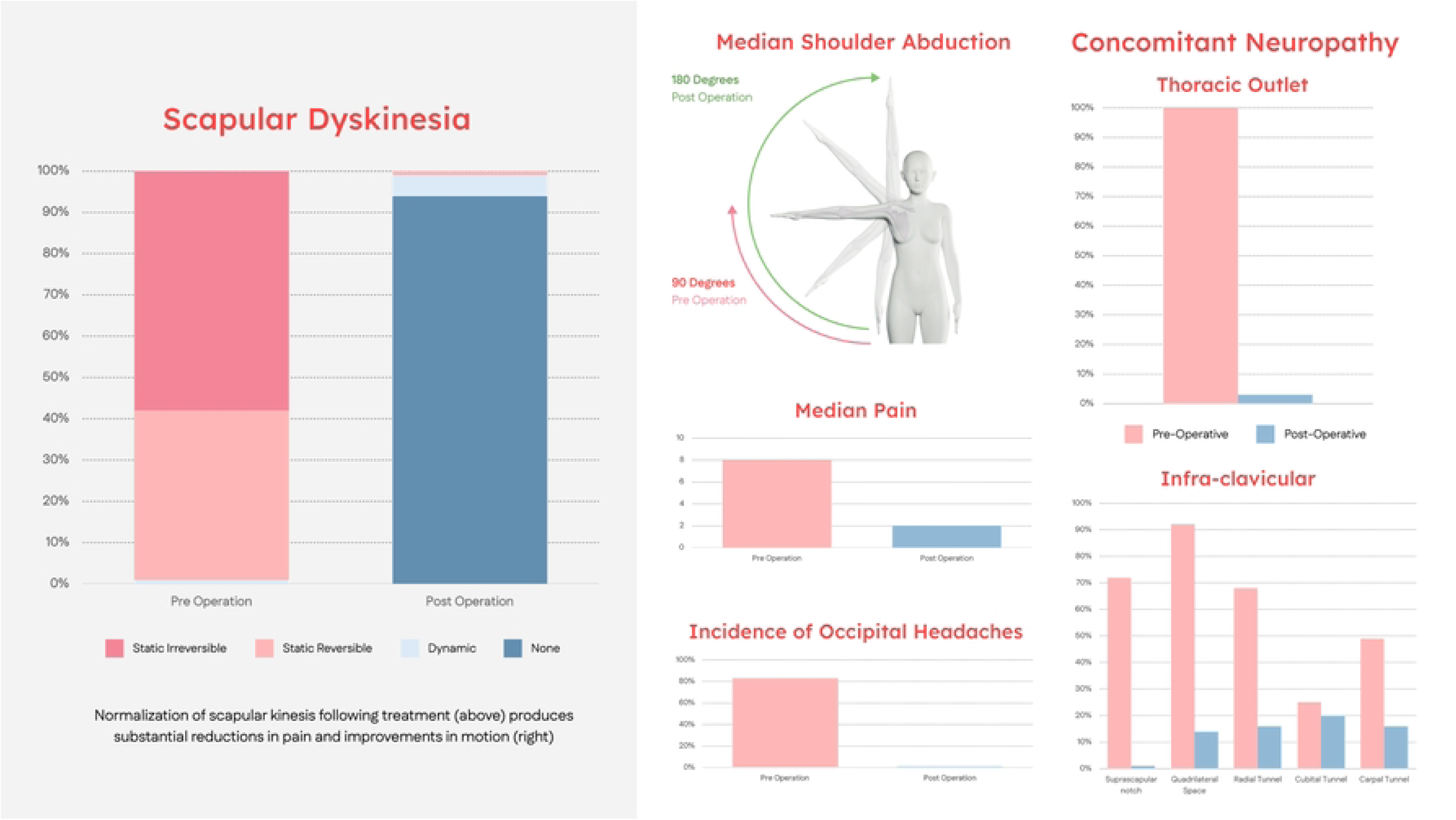
Normalization of Scapular Dyskinesis Resolves Symptoms in Thoracic Outlet Syndrome Following PM tenotomy with infraclavicular neurolysis alone for TOS, the normalization of the scapula mechanics reduces pain, neuropathy, and headaches and restores motion, suggesting that TOS is a scapular phenomenon.

**Figure 4.**
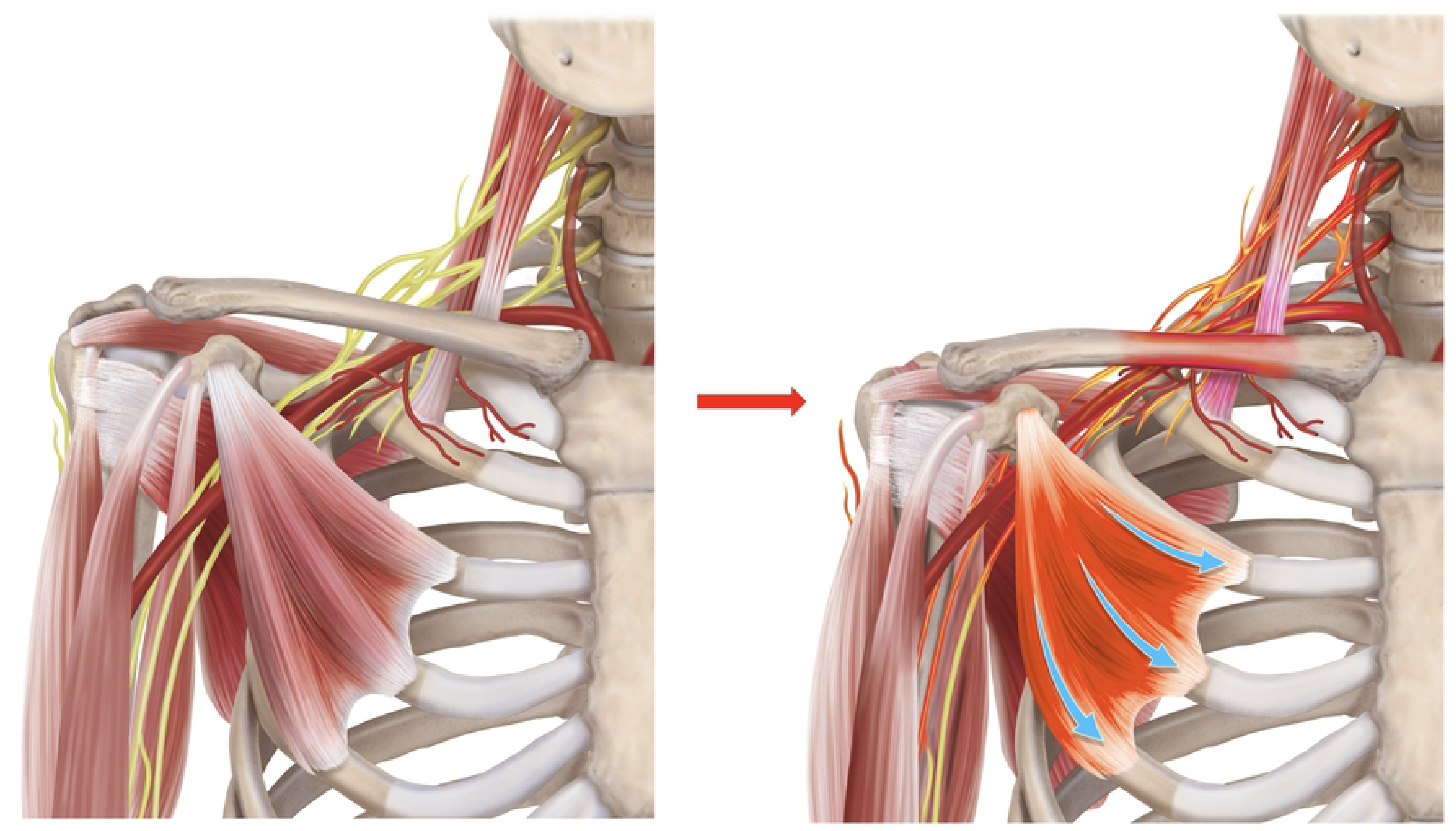
Pathoanatomy of How Human Disharmony Loop Produces Thoracic Outlet Syndrome Ventrally, the overpowering PM pull tugs the coracoid inferior and medial *(blue arrows)*. Consequently, the acromion and AC joint and therefore clavicle are pulled as well, narrowing the thoracic outlet. This compresses the subclavian vessels and brachial plexus and stretches the scalene muscles.

While TOS has always been considered compression of the brachial neurovascular bundle[7], many findings suggest this understanding is incomplete. These patients exhibit many symptoms outside the domain of neurovascular compression, including occipital headaches[27, 28], neck, upper back, peri-trapezial and peri-clavicular pain[12, 14, 25, 26], and chronically slumped postures.[4] Tellingly, the ipsilateral scapula protracts with dyskinetic mechanics, but explanations for this have been lacking.[4, 29, 30] If the etiology is truly compression, the questionable efficacy and high recurrence rate of surgeries which decompress the thoracic outlet[12, 13, 22, 24] is challenging to explain. Furthermore, without an anatomic basis to describe why the neurovascular compression occurs, a diverse basket of vague and unexplained “causes” must be invoked.[7, 8, 19, 26] The surgeries can be morbid with complications that span pneumothorax, chyle leak, vascular injury, and brachial plexus injury.[23] Another mystery is the preponderance towards women – replicated here – which similarly lacks convincing reasoning.[7, 32] All of these considerations complicate decision making for patients and practitioners.

In contrast, the HDL cleanly settles these points of contention. (Table 3) Fundamentally, the HDL is a scapular phenomenon, which includes but extends beyond neurovascular compression. The HDL articulates why TOS patients develop occipital headaches (occipital neuritis), neck and upper back pain (upper trapezius and rhomboid stretch), peri-clavicular pain (clavicle displacement), and chronically hunched shoulders (self-reinforcing PM pull).[34, 36] Crucially, the HDL resolves why the scapula protracts: scapula protraction causes TOS, not the other way around. The HDL also explains both the higher prevalence in women, as the weight of the breast tissue tightens the PM[38], and the lofty recurrence rate after surgical decompression of the thoracic outlet is also explained, as narrowing here is an effect and not the root cause. (Figure 1) In contrast to TOS which suffers from vague and unreliable diagnosis[13, 16, 18] and morbid surgeries[23], the HDL advances clear diagnostic criteria and offers an outpatient surgical treatment with high efficacy and low morbidity, conferring a <1% major complication risk.[34] Furthermore, the HDL can anatomically connect TOS to other concomitant pathologies, including subacromial impingement, cervical stenosis, and scapular dyskinesis.

**Table 3.** Key Differences between Conventional Thoracic Outlet Syndrome (TOS)/Pectoralis Minor Syndrome (PMS) versus Human Disharmony Loop (HDL)

|  | **<u>TOS/PMS</u>** | **<u>HDL</u>** |
| --- | --- | --- |
| Root Etiology | **Unknown:** tend to invoke numerous vague “causes” which themselves remain unexplained (“postural spasm”, “abnormal mechanics”, etc..) | **Singular:** unique asymmetric lower trunk innervation to PM |
| Mechanism | Neurovascular **compression** | Deformation of **scapula** and its connections |
| Diagnostic Criteria | Vague and unreliable | Clear and reproducible |
| Symptoms Explained by Proposed Etiology | Numbness/tingling, weakness, coolness only | **All chronic pain symptoms:** headaches, neck pain, shoulder weakness, myofascial trigger points, scapular protraction, slumped posture, and numbness/tingling, weakness, coolness |
| Anatomic Relationships to Other Chronic Pain Entities | **Unknown** | Clearly identifies **cause and effect** |
| Epidemiology | Considered rare | Ubiquitous |

Conventional TOS/PMS and the HDL are distinct clinical syndromes whose critical differences are highlighted here. In contrast to TOS/PMS, which is fundamentally a compressive neuropathy, HDL is a scapular phenomenon. Therefore, unlike TOS, the HDL can (1) explain the root etiology, (2) be diagnosed easily with clean and reproducible criteria, (3) account for all chronic pain symptoms seen and not just neuropathy, and (4) articulate clear cause and effect anatomic pathways to other known chronic pain entities.

To date, the PM has largely been considered incidental or secondary rather than causative in upper limb pathology. Conventional understanding claims the PM serves as an infra-clavicular compression point.[39, 40] In all these models, PM tightness or shortening develops secondarily after scapular dyskinesis[40] or trauma, repetitive stress, and overhead activity[11, 15, 41], and occurs simultaneously with TOS.[42] Specifically, “over time, a hyperactive or spasming PM shortens and develops contracture”[40] and “subcoracoid compression arises as a later secondary feature due to prolonged postural changes, chronic muscle spasm, and imbalances in shoulder girdle mechanics.”[26] However, how these vague abnormalities arise and why they lead to PM contracture remains unknown:[40, 43, 44] “the cause of PM tightness is not fully understood.”[45] In contrast, within the HDL, the PM is causative and central. PM tightness arises solely due to its unique lower trunk innervation, and then causes scapular dyskinesis[36], deforms the scapula’s numerous connections, and produces TOS, postural changes, chronic muscle spasm, imbalanced shoulder girdle mechanics, etc. These alleged “causes” are sequelae of the asymmetric neurologic innervation around the human scapula which predisposes the scapula to protraction (or the shoulder to hunching). Notably, 16% of patients in this study required subsequent secondary neurolysis, consistent with double-crush[4], highlighting the importance of surveying patients for residual neuropathy, but only 1% underwent SCN±FRR. We advocate a strategy of ‘breaking the loop’ first via PM+ICN, followed by close surveillance with secondary treatment for residual neuropathic lesions. Of note, three patients in this study had failed prior SCN+FRR but responded well to PM+ICN, reinforcing that the PM is causative in HDL patients.

This study suffers from key limitations. As a case series limited to two sites, our results must be replicated before widespread adoption. The lack of control group means unknown confounders and/or placebo effect could explain the findings. Neuropathy was diagnosed via the SCT which suffers drawbacks[46], but no gold standard exists.[47] Critically, our results only apply to TOS patients who meet HDL criteria, and future studies need to explore patients with TOS symptoms who are not ‘in the loop’. Most patients in this study are nTOS, and additional work is required to investigate the relationship between vTOS/aTOS to the HDL. Certainly, compression within the interscalene or costoclavicular spaces can occur independent of the HDL, so thorough assessment is critical to triage patients properly. Nonetheless, we do maintain that for patients who meet HDL criteria, PM+ICN with staged SCN±FRR if necessary is an appropriate strategy, given the 99% efficacy of the former and high morbidity of the latter. To definitively prove our assertions, future studies should randomize PM+ICN vs. SCN±FRR in large, multi-institutional trials, although this may be challenging given the heterogeneity of diagnosis and treatment for TOS. Our median follow-up of 15 months is not sufficient to prove permanence, and patients may regress over time.[24]

Nonetheless, in a large series of TOS patients, isolated PM+ICN significantly reduced pain, nearly eliminated headaches, and restored motion, with only 1% requiring subsequent SCN+FRR. The HDL anatomically explains all symptoms, answers vexing mysteries, and offers an effective and low-morbidity surgical alternative. We propose that TOS is fundamentally a scapular phenomenon; a subset of the HDL itself, the inherent instability of the human scapula to revert to its quadrupedal state.[33] Future research is required to confirm or refute these findings.

## Data Availability

https://figshare.com/articles/dataset/TOS_Dataset_Deidentified/30735830?file=60017951

